# Deaths induced by compassionate use of hydroxychloroquine during the first COVID-19 wave: the devil is in the details

**DOI:** 10.1101/2024.01.18.24301464

**Authors:** Charlotte Beaudart, Jonathan Douxfils, Flora Musuamba, Jean-Michel Dogné

**Affiliations:** Clinical Pharmacology and Toxicology Research Unit, Faculty of Medicine, NARILIS, Université de Namur, Namur, Belgium; QUALIresearch, QUALIblood s.a., Namur, Belgium; Department of Biological Hematology, Centre Hospitalier Universitaire Clermont-Ferrand, Hôpital Estaing, Clermont-Ferrand, France

## Abstract

Several trials of different designs were conducted to investigate the efficacy and safety of hydroxychloroquine (HCQ) for the prevention and/or the treatment of COVID-19 patients. Recently, it has been reported that HCQ might have been associated with an excess of 16,990 deaths during the first wave of the COVID-19 pandemic in 6 countries. Such attributable risk analysis is associated with many limitations. These previous findings did not adequately address dose-subgroup and sensitivity analyses which precludes any overall firm conclusions on in-hospital mortality attributable to HCQ.

We performed a meta-analysis and proposed a stratification by doses of HCQ. By pooling studies employing HCQ doses < or = 2400mg/5 days (i.e., k=12, n patients treated with HCQ=947, n controls=745), an OR of 0.94 (95%CI 0.56; 1.59) was found, indicating no increase in the mortality rate anymore. Importantly, there was no significant reduction in mortality rate with HCQ at < or = 2400mg/5 days neither. The same observation held true when pooling studies employing HCQ doses < or = 4800mg/5 days (i.e., k=25, n patients treated with HCQ=1672, n controls=1479) with an OR of 0.97 (95% CI 0.73; 1.29). Only high dose regimens of HCQ are associated with a significant increase in mortality.

Applying an excess of mortality in the population treated with doses where no increase of mortality is found creates a misleading overestimation of deaths associated with the use of HCQ in hospitalized patients with COVID-19. On the other hand, even at low doses HCQ regimen, no reduction in mortality with HCQ was observed suggesting that, when it comes to mortality as the outcome, HCQ did not show a benefit in hospitalized patients suffering from COVID-19. This mainly justifies the past and still up-to-date recommendations and guidelines to not use HCQ in this indication.

## Main text

Hydroxychloroquine (HCQ) used to treat malaria and autoimmune diseases such as rheumatoid arthritis gained significant attention in early 2020 as a potential treatment for COVID-19 based on its in vitro antiviral properties against SARS-CoV and SARS-CoV-2 and its favorable safety profile at recommended usual doses in acute and chronic use. HCQ was selected by the World Health Organization (WHO) and other medicines regulatory bodies for repurposing for COVID-19. For instance, in Belgium, off-label use of HCQ in monotherapy was recommended for hospitalized COVID-19 patients at a carefully selected dose based on limited pharmacokinetic models suggesting that a dosage of 400 mg twice daily for 1 day, followed by 200 mg twice daily for another 4 days (i.e. a total of 2,400 mg in total over 5 days) should have sufficient antiviral activity.[1, 2]

Several trials of different designs were conducted to investigate the efficacy and safety of HCQ for the prevention and/or the treatment of COVID-19 patients.[3] As the pandemic unfolded, conflicting findings emerged from clinical trials and observational studies. Some early studies indicated potential benefits, while others raised issues about the drug’s safety and efficacy.[4] Indeed, concerns about potential adverse drug reactions, including heart rhythm abnormalities and increased mortality,[5, 6] had prompted regulatory agencies to caution against its use outside of controlled settings.[6] The WHO and other health authorities reversed their recommendations of the use of HCQ in COVID-19 patients based on accumulating evidence.[7-9]

Several manuscripts aimed at gathering the information available in published and unpublished data to demystify these conflicting results and claims.[3, 6-9] Recently, Pradelle *et al*. estimated the in-hospital mortality attributable to HCQ during the first wave of COVID-19 by combining the mortality rate, HCQ exposure, number of hospitalized patients, and the increased relative risk of death with HCQ.[10] The main finding of their study is that HCQ might have been associated with an excess of 16,990 deaths during the first wave of the COVID-19 pandemic in the 6 countries for which data were available. Such attributable risk analysis is associated with many limitations, some of which being identified by the authors.[10] However, we want to point out the major limitation that their study did not adequately address dose-subgroup and sensitivity analyses which precludes any overall firm conclusions on in-hospital mortality attributable to HCQ.

The authors utilized the odds ratio (OR) published by Axfors *et al*., which encompasses 14 published and 15 unpublished trials as the estimator for HCQ-related mortality.[6] This meta-analysis reported an OR of 1.11 (95% CI 1.02; 1.20) and was based on 4,316 patients treated with HCQ and 5,696 controls. The outcomes reported by Pradelle *et al*. were heavily influenced by this effect size.[10] However, the significance of this effect size of 1.11 should be interpreted with caution and specific estimates based on the doses administered should be used. Indeed, two studies, namely WHO SOLIDARITY and RECOVERY, contribute to 88.9% of the weights in their overall model.[6] In essence, the pooled OR obtained from the meta-analysis is heavily influenced by these two specific trials. As highlighted by the authors themselves, RECOVERY and WHO SOLIDARITY employed HCQ in comparatively higher doses than all other trials, which may explain the increased OR observed while including them in the model.

To provide a nuanced analysis of the impact of this aspect on the overall results, we reiterated their meta-analysis and conducted a dose-subgroup analysis to assess whether using lower doses of HCQ (e.g., ≤2400mg/5 days or ≤4800mg/5 days) also significantly increased the risk of mortality across trials. As suggested above, the ‘low-dose’ HCQ regimen (2400 mg in total over 5 days) was recommended and used as a reasonable regimen for hospitalized patients.[1] Our analyses revealed that when pooling studies employing HCQ doses ≤2400mg/5 days (i.e., k=12, n patients treated with HCQ=947, n controls=745), an OR of 0.94 (95%CI 0.56; 1.59) was found (**Figure 1A**), indicating no increase in the mortality rate anymore. Importantly, there was no significant reduction in mortality rate with HCQ at ≤2400mg/5 days neither. The same observation held true when pooling studies employing HCQ doses ≤4800mg/5 days (i.e., k=25, n patients treated with HCQ=1672, n controls=1479) with an OR of 0.97 (95% CI 0.73; 1.29). Only high dose regimens of HCQ are associated with a significant increase in mortality (**Figure 1A & Figure 1B**).

**Figure 1.**
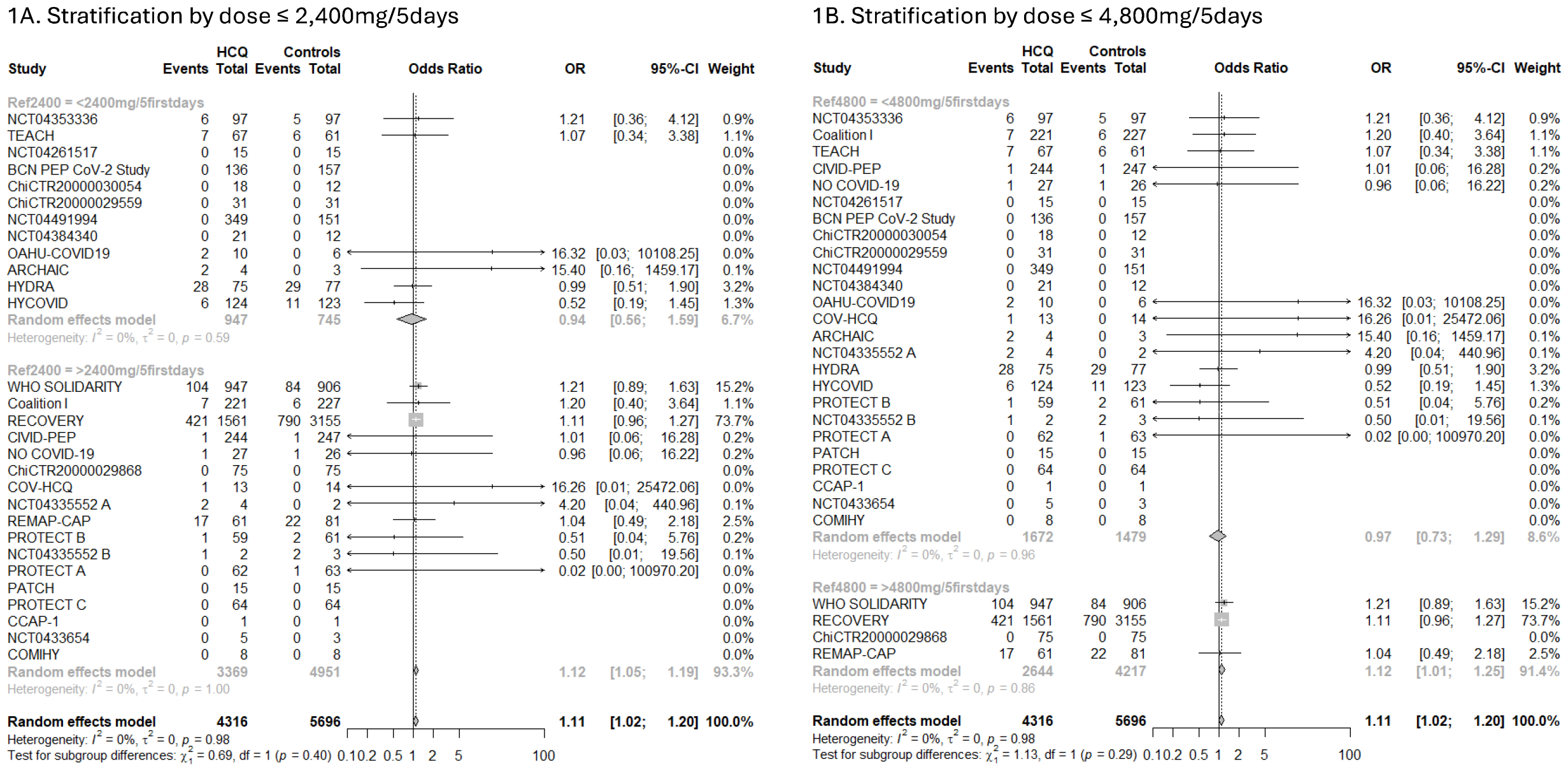
Random effect meta-analysis of mortality for the treatment of COVID-19 trials stratified by dose. **1A** Random effect meta-analysis of mortality for the treatment of COVID-19 trials stratified by dose (≤2400mg/5 days vs. >2400mg/5 days). **1B** Random effect meta-analysis of mortality for the treatment of COVID-19 trials stratified by dose (≤4800mg/5 days vs. >4800mg/5 days). The dose of HCQ received during the first 5 days of hospitalization was calculated from Table 2 of Axfors et al. presenting the group-level characteristics of each included randomized controlled trial. We used the random effect model of the Hartung-Knapp-Sidik-Jonkman (HKSJ) approach with the Paule-Mandel (PM) estimator for tau2, as it was the statistical method used by Axfors et al. All analyses were performed on R, version 2023.09.0+463, using the “meta” package. Scripts related to this analysis is freely available on Open Science Framework (https://osf.io/ewudy/).

Besides this methodological concern of applying an effect size found exclusively for high-dose studies to all patients, regardless of the dose they might have received, this OR of 1.11 has not been demonstrated to be robust. Indeed, Axfors *et al*.[6] did not conduct a leave-one-out analysis despite this sensitivity analysis is considered as a crucial methodological step to assess the robustness of a model. Interestingly, upon excluding either the WHO SOLIDARITY or the RECOVERY study from the model in leave-one-out analysis, the significance of the results is annulled (omitting WHO SOLIDARITY: OR 1.08 (95%CI:0.99; 1.19), omitting RECOVERY: OR 1.11 (95%CI:0.95; 1.30), plot available in Open Science Framework https://osf.io/ewudy/). The robustness of a meta-analytic model should be ensured through sensitivity analyses, and the significance of an effect size should not be attributable to solely one single trial. Furthermore, Axfors *et al*. ran additional sensitivity analyses to assess the robustness of their results across four different meta-analytic approaches (reported in their Appendix).[10] From these results, it is noteworthy that only one of the meta-analytic approaches tested (i.e. the Hartung-Knapp-Sidik-Jonkman (HKSJ) model with the Paule-Mandel estimator for tau2) yielded to a statistically significant OR of 1.11, while the three other statistical approaches failed to demonstrate the statistical significance of the effect size. When embarking on a study of such public health interest, Pradelle *et al*.[10] should have ensured that the main effect size on which they based their analysis,[6] and which was consistently employed across their models to estimate the number of excess deaths, was robust and unbiased. Our reanalysis points out that this is not the case, rendering their results unreliable.[10]

As a more general comment, the dosing regimen is critical in the development of new medicines and mainly for drug repurposing in the absence of already available robust clinical data. Underdosing and overdosing may lead to a lack of efficacy, and adverse drug reactions, respectively, which could potentially impact the net clinical benefit. Understanding the dose/concentration-dependent efficacy and toxicity of HCQ, as well as their determinants is therefore essential in assessing the risk-benefit trade-off in COVID-19 clinical trials. When a risk such as QTc prolongation is identified with HCQ, additional measures and warning should be implemented including QTc determination in all admitted patients and close cardiac monitoring to minimize such a risk.

In conclusion, applying an excess of mortality in the population treated with doses where no increase of mortality is found creates a misleading overestimation of deaths associated with the use of HCQ in hospitalized patients with COVID-19. On the other hand, even at low doses HCQ regimen, no reduction in mortality with HCQ was observed suggesting that, when it comes to mortality as the outcome, HCQ did not show a benefit in hospitalized patients suffering from COVID-19. This mainly justifies the past and still up-to-date recommendations and guidelines to not use HCQ in this indication.

## Data Availability

Material related to this analysis is freely available on Open Science Framework (https://osf.io/ewudy/).

## Declarations

### Author’s statement

All the authors approved the last version of the manuscript.

